# KNOWLEDGE AND COMPLIANCE TO OCCUPATIONAL RADIATION PROTECTION IN ENDOVASCULAR PROCEDURES: A NATIONAL CROSS-SECTIONAL SURVEY AMONG VASCULAR SURGEONS IN BRAZIL

**DOI:** 10.64898/2026.06.15.26355263

**Authors:** Andressa Cristina Sposato Louzada, Caroline de Aguiar dos Santos, Estevão Araujo Epifanio, Bruno Jeronimo Ponte, Carolina Carvalho Jansen Sorbello, Juliana Matsumura, Antonio Eduardo Zerati, Edwaldo Edner Joviliano, Nelson Wolosker

## Abstract

**Introduction:** The advent of Endovascular surgery and the use of fluoroscopy-guided procedures have grown in the last decade, and with that increased the exposure of surgeons to cumulative ionizing radiation, increasing the risk of occupational health complications. Despite established radiation protection principles, adherence to radioprotection measures and surveillance practices remains uncertain in many settings. This study aimed to evaluate knowledge, availability, and implementation of radiation protection strategies among vascular surgeons in Brazil.

**Metology:** A national cross-sectional survey was conducted in February 2026 using an anonymous online questionnaire distributed to all active members of the Brazilian Society of Angiology and Vascular Surgery (SBACV). Associations between participant characteristics and radioprotection practices were explored using chi-square or Fishers exact tests.

**Results:** Of 4,698 invited members, 192 vascular surgeons met the inclusion criteria. Most participants were male (67.4%), with a mean age of 45.9 years and a median of 12 years of experience performing fluoroscopy-guided procedures. Basic PPE use, particularly lead aprons, was nearly universal; however, adherence to other protective measures was substantially lower. Most respondents reported employing radiation-reduction strategies, including minimizing fluoroscopy time (98.9%), collimation (90.6%), optimized table and detector positioning (88.3%), procedural planning (87.2%), and pulsed fluoroscopy (81.0%). Only 19.4% reported undergoing annual medical surveillance for radiation-related health effects. Cataracts were the most frequently reported radiation-associated condition (6.2%). Greater age and professional experience were associated with higher utilization of selected protective measures and advanced imaging strategies.

**Conclusion:** The study provides important insights into how radiation protection is currently understood and implemented in contemporary Brazilian vascular surgery practice. The incomplete use of personal protective equipment (PPE), with many justifications in addition to the low surveillance of the effects of radiation on health, brings us an alert about the reality in Brazil.

## Introduction

The advent of endovascular surgery enabled faster postoperative recovery, shorter hospital stays, and potential improvements in overall care efficiency.(1–3) But its widespread adoption has been accompanied by a growing concern about the occupational hazards associated with ionizing radiation exposure. Cumulative exposure has been linked to an elevated risk of leukemia(4), brain tumors(5), thyroid and other solid organ malignancies(6), cataracts(7), accelerated atherosclerosis(8), dermatological injuries(9) and orthopedic injuries(10).

Complex endovascular aortic aneurysm repairs are among the fluoroscopy-guided procedures (FGP) associated with the highest radiation output(9), given the extensive angiographic acquisition required (8) and the inherent duration and technical complexity of these procedures.(11) In such cases, the dose received in a single procedure may reach the annual limit considered safe, with the hands and eyes being the areas most affected.(12) Accordingly, endovascular surgeons are formally classified as Category A occupational radiation workers, a designation that mandates structured medical surveillance for radiation-related health effects(4).

All professionals performing FGPs are required to adhere to the ALARA principle — As Low As Reasonably Achievable — which advocates for minimizing radiation exposure while preserving diagnostic image quality, thereby ensuring both procedural safety and clinical efficacy.(13) Despite this established framework, recent evidence suggests that a substantial proportion of professionals who perform endovascular procedures lack adequate knowledge of radiation exposure risks and radioprotection standards (14–17). Furthermore, the availability of protective resources and shielding equipment varies considerably across institutions and healthcare settings.

To the best of our knowledge, only a few national studies addressing this important issue. Two studies were conducted in the United States and evaluated only vascular trainees (16, 17) and two evaluated both vascular trainees and consultants in Spain and Great Britain plus Ireland (15,19). All four studies identified relevant gaps in the implementation of ALARA strategies.

Considering these findings, the present study was designed to assess the extent to which professionals performing FGPs adhere to individual radiation protection measures and the ALARA principles in Brazil, and to identify the underlying reasons for non-compliance where applicable.

## Materials and Methods

A cross-sectional, descriptive, and quantitative study was conducted in February 2026 using an anonymous, self-administered online questionnaire that could be completed only once. The survey instrument was hosted on the REDCap (Research Electronic Data Capture) platform for two-weeks. The study was approved by the Institutional Review Board under protocol number 6.771.348.

An invitation to participate in the study was distributed via email by the Brazilian Society of Angiology and Vascular Surgery (SBACV) to all active members. Participants who accepted the invitation, accessed the questionnaire, completed the informed consent form, formally agreeing to participate in the research, and answered more than 50% of the questions asked were included in the study. Responses were collected anonymously, and no personally identifiable information was obtained from participants.

Data collected included sociodemographic characteristics, professional practice information, and adherence to ALARA principles, including the habitual use of individual and collective protective equipment. Additionally, the self-reported history of conditions potentially related to occupational exposure to ionizing radiation was assessed, as well as the participants’ knowledge of possible complications resulting from chronic and cumulative exposure, including cataracts, thyroid disorders, neoplasms, dermatological lesions, and cardiovascular effects.

Exploratory analyses were performed to investigate potential associations between participants’ professional and institutional characteristics and the use of radiation protection measures. Associations between resource utilization and both years of professional practice and age were assessed by categorizing participants according to the median years of practice (12 years) and median age (43.5 years). Furthermore, associations between the use of protective equipment and procedural resources were evaluated by sex and type of institution (public, private, or both).

### Statistical Analysis

Categorical variables were summarized using absolute and relative frequencies. Continuous variables were described using measures of central tendency and dispersion, including means and standard deviations (SDs), medians and interquartile ranges (IQRs), and minimum and maximum observed values. When appropriate, results were also presented graphically using bar charts to facilitate data visualization and interpretation.

Associations between categorical variables were assessed using the chi-square or Fisher’s exact test, as appropriate, based on the characteristics of the variables and the distribution of the observed data. All statistical analyses were performed using the R software (R Foundation for Statistical Computing, Vienna, Austria). A two-sided p-value < 0,05 was considered statistically significant.

## Results

Of the 4,698 active SBACV members invited to participate in the study, 192 (4.1%) completed the questionnaire and met the eligibility criteria for inclusion.

Table 1 summarizes the sociodemographic and professional characteristics of the participants.

**Table 1.**
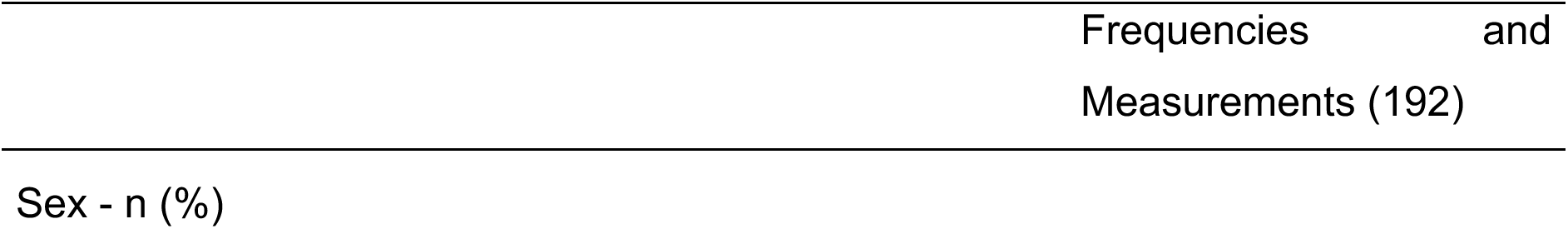
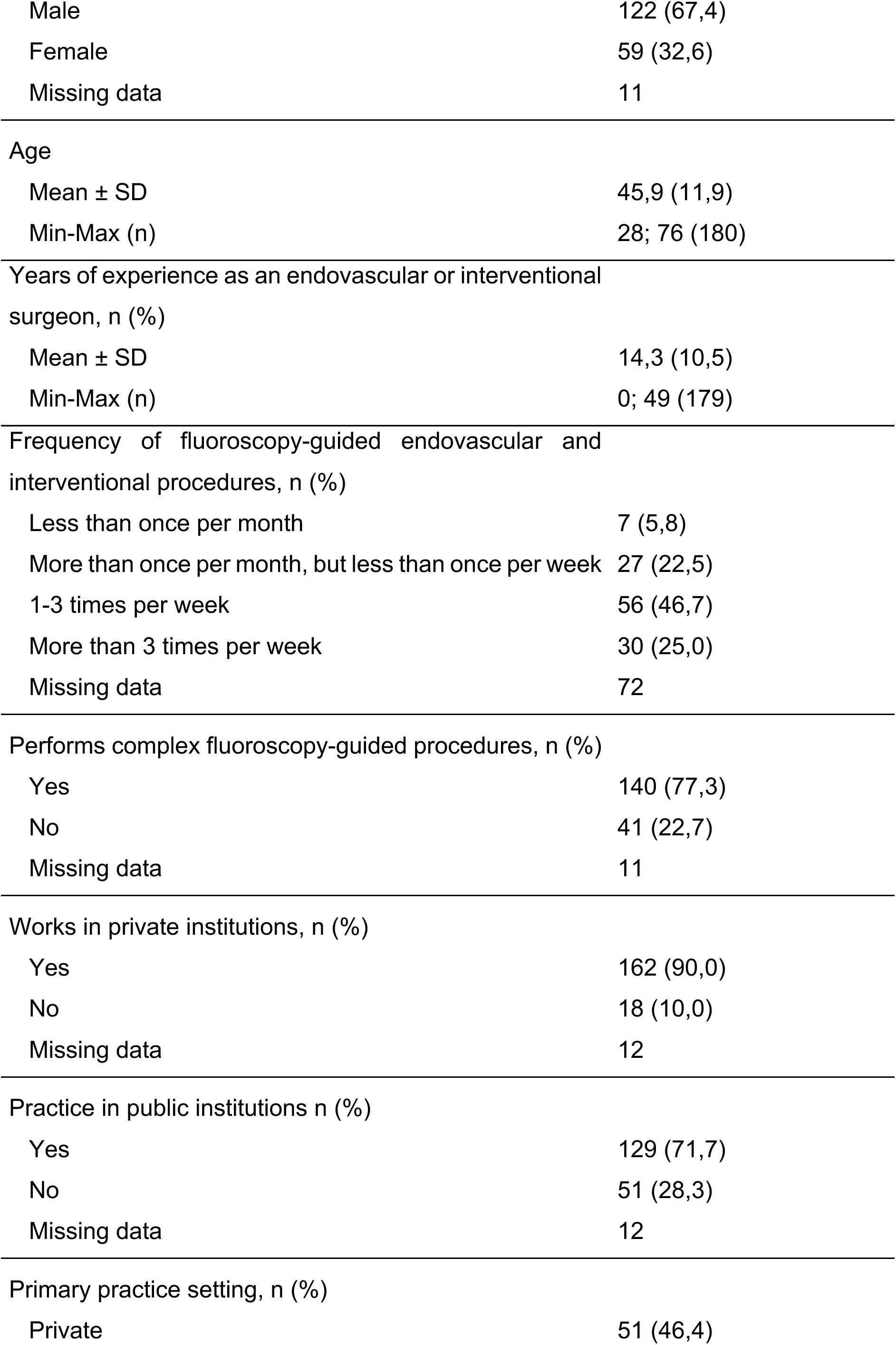
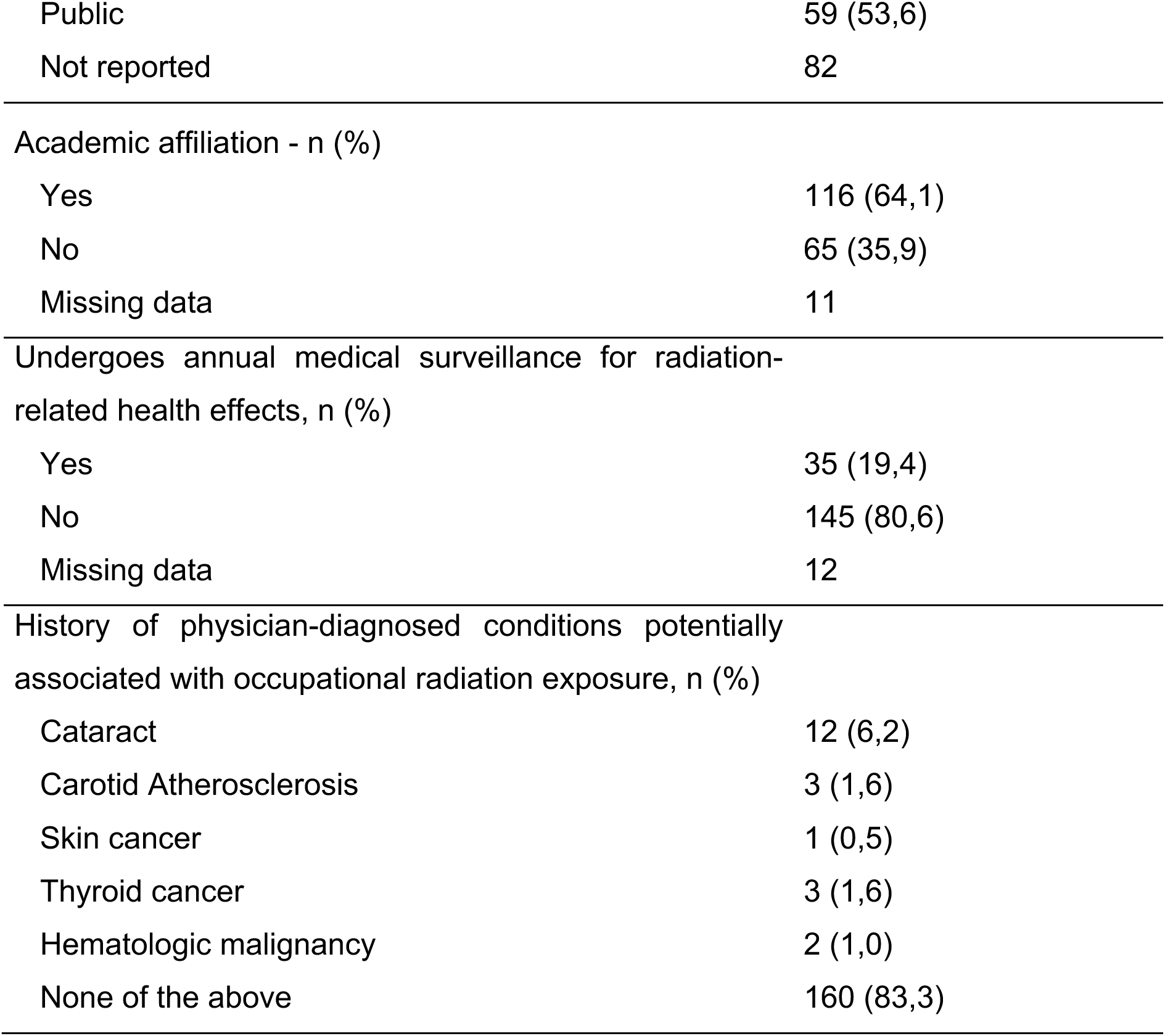
Sociodemographic and professional characteristics, clinical history related to occupational radiation exposure.

|  | Frequencies and<br>Measurements (192) |
| --- | --- |
| Sex - n (%) |  |
| Male | 122 (67,4) |
| Female | 59 (32,6) |
| Missing data | 11 |
| Age |  |
| Mean ± SD | 45,9 (11,9) |
| Min-Max (n) | 28; 76 (180) |
| Years of experience as an endovascular or interventional surgeon, n (%) |  |
| Mean ± SD | 14,3 (10,5) |
| Min-Max (n) | 0; 49 (179) |
| Frequency of fluoroscopy-guided endovascular and interventional procedures, n (%) |  |
| Less than once per month | 7 (5,8) |
| More than once per month, but less than once per week | 27 (22,5) |
| 1-3 times per week | 56 (46,7) |
| More than 3 times per week | 30 (25,0) |
| Missing data | 72 |
| Performs complex fluoroscopy-guided procedures, n (%) |  |
| Yes | 140 (77,3) |
| No | 41 (22,7) |
| Missing data | 11 |
| Works in private institutions, n (%) |  |
| Yes | 162 (90,0) |
| No | 18 (10,0) |
| Missing data | 12 |
| Practice in public institutions n (%) |  |
| Yes | 129 (71,7) |
| No | 51 (28,3) |
| Missing data | 12 |
| Primary practice setting, n (%) |  |
| Private | 51 (46,4) |
| Public | 59 (53,6) |
| Not reported | 82 |
| Academic affiliation - n (%) |  |
| Yes | 116 (64,1) |
| No | 65 (35,9) |
| Missing data | 11 |
| Undergoes annual medical surveillance for radiation-related health effects, n (%) |  |
| Yes | 35 (19,4) |
| No | 145 (80,6) |
| Missing data | 12 |
| History of physician-diagnosed conditions potentially associated with occupational radiation exposure, n (%) |  |
| Cataract | 12 (6,2) |
| Carotid Atherosclerosis | 3 (1,6) |
| Skin cancer | 1 (0,5) |
| Thyroid cancer | 3 (1,6) |
| Hematologic malignancy | 2 (1,0) |
| None of the above | 160 (83,3) |

|  | Frequencies and Measurements (192) |
| --- | --- |
| Sex - n (%) |  |
| Male | 122 (67,4) |
| Female | 59 (32,6) |
| Missing data | 11 |
| Age |  |
| Mean ± SD | 45,9 (11,9) |
| Min-Max (n) | 28; 76 (180) |
| Years of experience as an endovascular or interventional surgeon, n (%) |  |
| Mean ± SD | 14,3 (10,5) |
| Min-Max (n) | 0; 49 (179) |
| Frequency of fluoroscopy-guided endovascular and interventional procedures, n (%) |  |
| Less than once per month | 7 (5,8) |
| More than once per month, but less than once per week | 27 (22,5) |
| 1-3 times per week | 56 (46,7) |
| More than 3 times per week | 30 (25,0) |
| Missing data | 72 |
| Performs complex fluoroscopy-guided procedures, n (%) |  |
| Yes | 140 (77,3) |
| No | 41 (22,7) |
| Missing data | 11 |
| Works in private institutions, n (%) |  |
| Yes | 162 (90,0) |
| No | 18 (10,0) |
| Missing data | 12 |
| Practice in public institutions n (%) |  |
| Yes | 129 (71,7) |
| No | 51 (28,3) |
| Missing data | 12 |
| Primary practice setting, n (%) |  |
| Private | 51 (46,4) |
| Public | 59 (53,6) |
| Not reported | 82 |
| Academic affiliation - n (%) |  |
| Yes | 116 (64,1) |
| No | 65 (35,9) |
| Missing data | 11 |
| Undergoes annual medical surveillance for radiation-related health effects, n (%) |  |
| Yes | 35 (19,4) |
| No | 145 (80,6) |
| Missing data | 12 |
| History of physician-diagnosed conditions potentially associated with occupational radiation exposure, n (%) |  |
| Cataract | 12 (6,2) |
| Carotid Atherosclerosis | 3 (1,6) |
| Skin cancer | 1 (0,5) |
| Thyroid cancer | 3 (1,6) |
| Hematologic malignancy | 2 (1,0) |
| None of the above | 160 (83,3) |

Most respondents were male (67.4%), with a mean age of 45.9 years. The median length of experience in fluoroscopy-guided procedures was 12 years. Regarding procedural volume, 46.7% reported performing endovascular procedures one to three times per week. Additionally, most respondents (77.7%) reported routinely participating in complex fluoroscopy-guided procedures.

Regarding practice setting, 90% of participants reported working in private institutions, while 71.7% also worked in public institutions. However, 53.6% reported that most of their professional activities were conducted in the public sector. Furthermore, 64.1% reported being affiliated with educational institutions.

### Health History

Only 19,4% reported having annual medical checkups for radiation-induced injury screening.

Among the conditions potentially associated with occupational exposure to ionizing radiation, cataracts were the most frequently reported, affecting 12 participants (6.2%). Carotid atherosclerosis and thyroid cancer were each reported by three participants (1.6%). Overall, 83.3% of respondents reported none of the evaluated conditions.

### Use of PPE during the procedure

Figure 1 illustrates the frequency of use of personal protective equipment (PPE) and the radiation-reduction strategies during fluoroscopy-guided procedures, as well as the reasons reported for their non-use.

**Figure 1.**
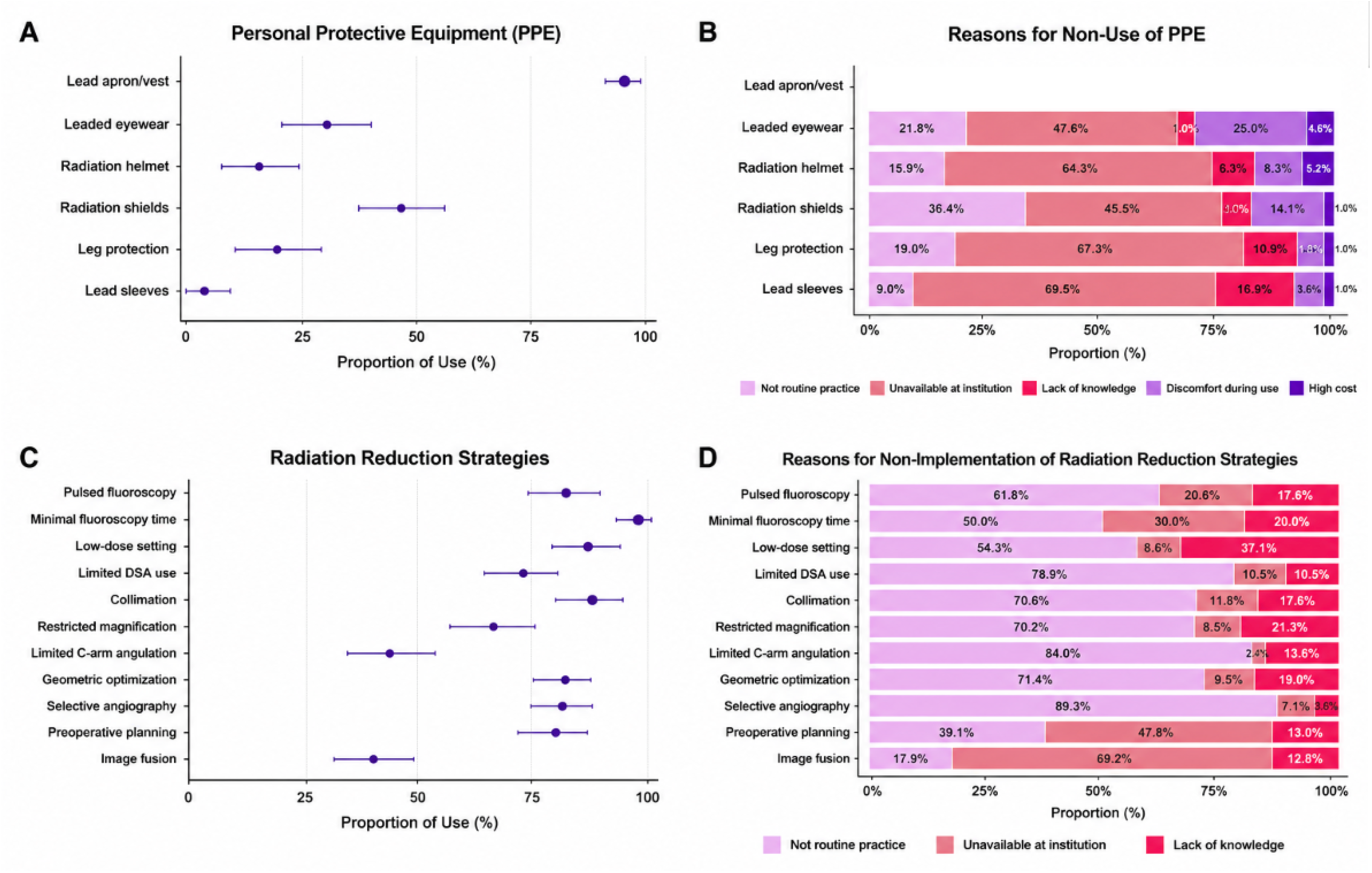
Proportion of use of protective equipment in fluoroscopy-guided procedures and reasons for non-use: *Panel “A” shows the proportion of PPE in fluoroscopy-guided procedures, and panel “B” presents the reasons used to justify not using PPE. Panel “C” illustrates the resources used to reduce exposure to radiation, and panel “D” summarizes the reason reported for non-implementation*.

The use of basic protective devices was nearly universal, particularly lead aprons or vests. In contrast, adherence to other radiation protection measures was substantially lower, including leaded eyewear, radiation shields, lower limb protection, helmets, and lead sleeves.

Among participants who did not use certain equipment, lack of availability within their institutions was the most frequently cited reason, especially for less commonly available or higher-cost equipment. Discomfort during use and the perception of lack of need were also mentioned, although less frequently.

### Image Resources

Most participants reported adopting strategies to reduce radiation exposure during procedures, such as limiting fluoroscopy pedal activation to the minimum time necessary (98.9%), collimation (90.6%), appropriate table and detector positioning (88.3%), preoperative procedural planning (87.2%), and the use of pulsed acquisition (81%). A substantial proportion of respondents also reported limiting the use of digital subtraction, restricting magnification, and minimizing C-arm angulation.

Among participants who did not employ these radiation-reduction measures, the most frequently cited reasons were lack of routine practice and the unavailability of the technology or resource in the service (Figure 1).

Age of vascular surgeons was significantly associated with greater adherence to the use of protective eyewear (p=0.048), pulsed fluoroscopy mode (p=0.043), and the use of image fusion technology (p=0.031) compared with their younger counterparts.

On the other hand, sex-based comparisons revealed significant differences in the use of certain protective devices. Male surgeons reported higher utilization of radiation shields than female surgeons (58,3% vs. 34,5%; p=0.005), as well as greater use of lower-extremity protectors (23,3% vs. 6,9%; p=0.014).

When evaluating the relationship between the time spent performing fluoroscopy and the use of imaging resources, comparing more experienced surgeons (>12 years of experience, median value) with those with less experience (<12 years), it was observed that the use of advanced image acquisition resources with image fusion was significantly more frequent among the more experienced (44% vs. 24.7%; p=0.011). Similarly, the use of pulsed fluoroscopy was more frequent among the more experienced surgeons (88.1% versus 74.2%; p=0.031). No significant associations were observed for the other imaging resources evaluated.

When comparing public and private institutions, a statistically significant association was observed between working in private institutions and the greater use of certain resources to reduce radiation exposure. Angle restriction was reported by 62.7% of professionals affiliated with private institutions, compared to 40.6% of those working in public institutions (p=0.034). Similarly, angiography was more frequently performed by the primary surgeon in private institutions (92.7% versus 74.6%; p=0.029). Among surgeons affiliated with teaching institutions, 57.4% reported the use of radiation shields (p=0.021) and 80% reported that angiography acquisitions were performed by the primary surgeon (p=0.048).

## Discussion

Ionizing radiation is widely employed to guide endovascular procedures, which are minimally invasive and yield excellent clinical results after conservative treatment. However, occupational exposure to ionizing radiation may harm both patients and healthcare professionals, particularly when exposure is repetitive and prolonged over the course of a career. Accordingly, its use must be governed by caution, technical responsibility, and rigorous adoption of available radioprotection measures.(16)

In this context, we conducted an innovative descriptive, quantitative, cross-sectional survey designed to assess knowledge, radioprotection practices, availability of personal protective equipment (PPE), and risk perception among Brazilian vascular surgeons — a population that has not been previously investigated.

The response rate was 4.1% of the total members invited. Although low, this figure must be interpreted within the context of a voluntary, anonymous online questionnaire distributed via email to a national medical society. And despite this response rate, our study has the largest sample size reported in the literature to date, comprising 192 endovascular surgeons.

The sample was predominantly composed of male vascular surgeons, with a mean age of 45.9 years and a median of 12 years of experience performing FGP. We observed that more experienced and senior surgeons were more likely to employ imaging strategies associated with lower radiation emission and scatter, and reported greater use of protective eyewear, suggesting heightened occupational risk awareness among those with greater cumulative exposure. (18) The finding of lower PPE adherence among younger surgeons has been similarly reported by other authors,(19) and educational gaps regarding ALARA principles among junior surgeons have also been documented in the literature. (16)

In addition to that, it drew our attention to the fact that more than 10% of the participants reported a lack of knowledge of technical strategies for reducing radiation exposure. All professionals should be trained in radioprotection principles early in their careers.(20) There is therefore an urgent need for institutions to promote periodic training programs grounded in ALARA principles,(21,22) with particular emphasis on younger practitioners.

Most of the participants (90%) reported working in private institutions and 71.7% in public institutions, an overlap that is highly prevalent in Brazilian medical practice. The majority reported a predominance of activity in the public sector, which is of particularly relevant given that the availability of protective equipment, institutional protocols, and occupational monitoring may vary substantially across different healthcare settings.

The most frequently cited reason for non-adherence to PPE was its unavailability in the workplace, a finding that was more prevalent in the public sector. Infrastructure, technological availability, workflow organization, and institutional investment capacity directly influence radioprotection practice. Our results suggest that the public sector faces greater structural limitations and more restricted access to protective resources, reinforcing the need for greater regulatory standardization across both systems, including the definition of mandatory minimum requirements to ensure the safety of surgeons, staff and patients.(23)

Among the 64.1% of participants affiliated with academic institutions, greater use of radiation shields was observed, and angiography was more frequently performed by the primary surgeon, suggesting an association between academic practice and higher adherence to certain protective measures — a finding consistent with those reported by other authors.(23,24)

Most technical dose-reduction strategies were widely reported, including minimizing fluoroscopy time, employing collimation, and optimizing table and detector positioning. However, the incomplete adoption of more advanced or infrastructure-dependent measures suggests that, despite of apparent familiarity with fundamental radioprotection principles and a reasonable level of operational awareness, surgeons remain vulnerable due to insufficient resources and infrastructure.(25)

Reports of cataracts, carotid atherosclerosis, and thyroid cancer must be interpreted with caution, as the cross-sectional, self-reported design of this study precludes the establishment of causal relationships. (26) Nevertheless, the prevalence of cataracts in 6.2% of participants is a noteworthy finding, given the well-documented radiosensitivity of the crystalline lens(12) and the fact that only 31.6% of respondents reported using lead-lined glasses — mainly due to their unavailability in their practice settings.

Furthermore, it is notable that only 19.4% of participants reported undergoing annual medical consultations for radiation-induced injury screening. This may reflect limited individual risk perception, the absence of clear institutional protocols, restricted access to occupational health programs, or the normalization of radiation exposure as an inevitable component of professional practice.(21) Given that international recommendations for Category A radiation workers mandate annual medical follow-up — in light of the potential for both stochastic effects, such as malignancy, and deterministic effects, such as cataracts(25) — there is an urgent need for institutions to implement structured screening protocols. This administrative disconnection and lack of systematic monitoring has been identified by other authors as well (Bhinder et al., 2022; Sritharan et al., 2024) and must be actively addressed by representative professional societies.

Thus, although most participants did not report conditions attributable to radiation exposure, the data reveal an important contradiction: this is a population frequently exposed to ionizing radiation, but who face a lack of regular medical check-ups and insufficient access to PPE. This scenario reflects a structural institutional failure and underscores the need to strengthen radioprotection programs, occupational monitoring systems, and continuing medical education among Brazilian vascular surgeons.(20,27)

Radiation protection remains a multifactorial challenge, encompassing professional behavior, ergonomics, resource availability, and organizational responsibility. Accordingly, periodic training programs have the potential to improve the standardization of practices and reduce inter-institutional variability.(28)

In summary, our findings indicate that most endovascular surgeons are familiar with and employ technical strategies to minimize ionizing radiation exposure; however, they often lack access to the PPE and protective resources needed to fully mitigate and monitor occupational hazards. These findings support the urgent need for more rigorous institutional policies to protect professionals involved in FGP.

## Limitations

This study has limitations, mainly related to the recruitment strategy, resulting in a low response rate of 4.1%. Furthermore, given that most participants reported performing complex endovascular procedures and nearly half reported performing FGP one to three times per week, selection bias cannot be excluded: endovascular surgeons with greater occupational radiation exposure were more likely to respond to this survey.

Nevertheless, our study achieved the largest number of respondents among comparable published investigations (Bhinder et al., 2022; Sritharan et al., 2024; Bordoli et al., 2014; Martínez Del Carmen et al., 2026) and incorporated strategies to reduce these limitations. The invitation was distributed by the SBACV to all active members, ensuring national reach and avoiding initial restriction to a limited number of institutions, regions, or high-complexity centers. This approach enabled the capture of perspectives from professionals working across diverse healthcare settings, including public, private, academic, and non-academic environments. Although participation was voluntary, the study provides meaningful insight into how radioprotection is currently understood and practiced in contemporary Brazilian vascular surgery.

Additionally, data were self-reported and therefore subject to recall bias and response imprecision. The use of multiple-choice questions, though necessary for instrument standardization, may not fully capture the complexity of individual practices.

Finally, this study was limited by the absence of detailed stratification by procedure type. Different endovascular interventions are associated with markedly different radiation exposure levels, which may limit the precision with which the relationship between occupational exposure and radioprotection adherence can be analyzed.

## Conclusion

The present study offers valuable insights into the current understanding and implementation of radiation protection practices among endovascular surgeons in Brazil. Among those reporting low adherence to personal protective equipment and the ALARA principles, the predominant justification was the inadequacy or unavailability of appropriate resources in their clinical settings. Notably, over 10% of respondents acknowledged insufficient knowledge of technical strategies for radiation dose reduction, and fewer than 20% reported undergoing medical surveillance for radiation-related health effects. Collectively, these findings shed light on the concerning occupational realities faced by endovascular surgeons in Brazil, underscoring an urgent need for structured educational initiatives and enhanced institutional measures to safeguard these professionals.

## Data Availability

All data produced in the present work are contained in the manuscript

